# Detection of Major Depressive Disorder Using Vocal Acoustic Analysis and Machine Learning

**DOI:** 10.1101/2020.06.23.20138651

**Authors:** Caroline Wanderley Espinola, Juliana Carneiro Gomes, Jessiane Mônica Silva Pereira, Wellington Pinheiro dos Santos

**Affiliations:** Departamento de Engenharia Biomédica, Universidade Federal de Pernambuco, Recife, Brazil; Hospital Ulisses Pernambucano, Recife, Brazil; Núcleo de Engenharia da Computação, Escola Politécnica da Universidade de Pernambuco, Recife, Brazil

**Author notes:** Corresponding author: Wellington Pinheiro dos Santos.

**Keywords:** Major depressive disorder, diagnosis, voice, acoustic parameters, machine learning, support vector machines

## Abstract

**Purpose:** Diagnosis and treatment in psychiatry are still highly dependent on reports from patients and on clinician judgement. This fact makes them prone to memory and subjectivity biases. As for other medical fields, where objective biomarkers are available, there has been an increasing interest in the development of such tools in psychiatry. To this end, vocal acoustic parameters have been recently studied as possible objective biomarkers, instead of otherwise invasive and costly methods. Patients suffering from different mental disorders, such as major depressive disorder (MDD), may present with alterations of speech. These can be described as uninteresting, monotonous and spiritless speech, low voice.

**Methods:** Thirty-three individuals (11 males) over 18 years old were selected, 22 of which being previously diagnosed with MDD, and 11 healthy controls. Their speech was recorded in naturalistic settings, during a routine medical evaluation for psychiatric patients, and in different environments for healthy controls. Voices from third parties were removed. The recordings were submitted to to a vocal feature extraction algorithm, and to different machine learning classification techniques.

**Results:** The results showed that support vector machines (SVM) models provided the greatest classification performances for different kernels, with PUK kernel providing accuracy of 89.14% for the detection of MDD.

**Conclusion:** The use of machine learning classifiers with vocal acoustics features has shown to be very promising for the detection of major depressive disorder, but further tests with a larger sample will be necessary to validate our findings.

## 1. Introduction

Clinical assessment and treatment in psychiatry currently depend on diagnostic criteria built entirely on expert consensus, instead of relying on objective biomarkers (Bzdok & Meyer-lindenberg, 2018). Such criteria, described in the Diagnostic and Statistical Manual, 5^th^ Edition (DSM-5), and in the International Classification of Diseases (ICD-10), are still considered the gold-standard for diagnosis in psychiatry (American Psychiatric Association, 2013). Nevertheless, those diagnostic systems have been criticized due to their absence of clinical predictability and neurological validity (Bzdok & Meyer-lindenberg, 2018), and their poor diagnostic stability (Baca-Garcia et al., 2007). While other medical fields hold markers of disease presence and severity, such as tumor volume measurement and biochemical blood tests, psychiatry still lacks routine objective tests (Bedi et al., 2015; Mundt, Vogel, Feltner, & Lenderking, 2012).

Historically, evaluation and treatment in psychiatry are based on reports from patients and on clinical evaluation (Mundt, Snyder, Cannizzaro, Chappie, & Geralts, 2007). Thus diagnosis and therapeutic decision are extremely sensitive to memory and subjectivity biases (Jiang et al., 2018). Considering this, over the last decades there has been an intense search for biomarkers for diagnosis and follow-up of psychiatric patients (Iwabuchi, Liddle, & Palaniyappan, 2013; Mundt et al., 2012), most of those being expensive and invasive (Higuchi, Tokuno, Nakamura, & Shinohara, 2018). Despite all efforts, instruments for assessment of mental disorders still remain a conundrum (Mundt et al., 2007).

Major depressive disorder (depression) is the most common mental disorder, affecting more than 300 million people worldwide (Sadock, Sadock, & Ruiz, 2017; World Health Organization, 2018). It is also a leading cause of disability and economic burden (Mundt et al., 2007, 2012). The global prevalence of depression was estimated to be 4.4% in 2015 (World Health Organization, 2017), with more women affected than men in a 2:1 ratio (Weinberger et al., 2017). In Brazil the prevalence of depression is currently the fifth largest in the world, 5.8% (World Health Organization, 2017), while its lifetime prevalence can be as high as 16.8% (Miguel, Gentil, & Gattaz, 2011).

Patients suffering from depression may present with low mood, irritability, anhedonia, fatigue, psychomotor retardation, cognitive impairment (difficulty in decision making, poor concentration) and disturbances of somatic functions (insomnia or hypersomnia, appetite disorders, changes in body weight). These symptoms are associated with intense suffering and decline in functioning, and may ultimately lead to suicide (American Psychiatric Association, 2013). Depression is associated with approximately half of all suicides globally (Cummins et al., 2015).

Early depressive symptoms such as psychomotor retardation and cognitive impairment are frequently related to disturbances in speech (Hashim, Wilkes, Salomon, Meggs, & France, 2016). Actually, patterns within depressed speech have been documented years ago (Mundt et al., 2007). In particular, the persistent altered emotional state in depression may affect vocal acoustic properties. As a result, depressive speech has been described by clinicians as monotonous, uninteresting and without energy. These differences could provide the detection of depression through analysis of vocal acoustics of depressed patients (Jiang et al., 2018).

Machine learning is an intensive field of research, with successful applications to solve several problems in health sciences, like breast cancer diagnosis (Cordeiro, Lima, Silva-Filho, & Santos, 2012; Cordeiro, Santos, & Silva-Filho, 2017; Cordeiro, Santos, & Silva-Filho, 2016; Cordeiro, Santos, & Silva-Filho, 2013; de Lima, S. M., da Silva-Filho, A. G., & dos Santos, 2016; Rodrigues et al., 2019; Azevedo et al., 2015; de Santana et al., 2018), Alzheimer’s disease diagnosis support based on neuroanatomical features (dos Santos, de Assis, de Souza, Mendes, & de Souza Monteiro, H. S., Alves, 2009; dos Santos, de Assis, de Souza, & Santos Filho, 2008; dos Santos, de Souza, & dos Santos Filho, 2007; Cruz, T., Cruz, T., & Santos, W., 2018), multiple sclerosis diagnosis (Commowick, O., Istace, A., Kain, M., Laurent, B., Leray, F. & M., … & Kerbrat, 2018), and many applied neuroscience solutions (da Silva Júnior et al, 2019; de Freitas et al., 2019).

Qualitative changes in speech from people suffering with depression have been reported decades ago (Darby & Hollien, 1977), e.g., reduction in pitch range (Vanello et al., 2012), increased number of pauses (Mundt et al., 2012), slower speech (Faurholt-Jepsen et al., 2016), and reduced intensity or loudness (Hönig, Batliner, Nöth, Schnieder, & Krajewski, 2014).

For the recognition of changes in mood state, prosodic, phonetic and spectral aspects of voice are relevant, in particular fundamental frequency (F0) or pitch, intensity, rhythm, speed, jitter, shimmer, energy distribution between formants, and cepstral features. Among those features, jitter is considered important for mood state recognition due to its ability to identify rapid temporary changes in voice (Maxhuni et al., 2016). Alteration of mel frequency cepstral coefficients (MFCC) are also found in depressed individuals. MFCCs consist of parametrical representation of the speech signal (Hasan, Jamil, Rabbani, & Rahman, 2004) and have been extensively studied as possible features for the detection of major depressive disorder (Cummins, Epps, Sethu, & Krajewski, 2014; Jiang et al., 2018).

In a sample of 57 depressed patients, Cohn et al. (2009) analyzed prosodic and facial expression elements using two machine learning classifiers: support vector machines (SVM) and logistic regression. Their accuracy for the identification of depression was 79-88% for facial expressions, and 79% for prosodic features.

In a study with adolescents, Ooi, Lech, & Brian Allen (2013) used glottal, prosodic and spectral features, and Teager energy operator for the prediction of early symptoms of depression in that age group, and reported accuracy of 73% (sensibility: 79%; specificity: 67%). In another study with a larger sample of adolescents, Low, Maddage, Lech, Sheeber, & Allen (2011) utilized the above attributes with the addition of cepstral parameters, and submitted to SVM and Gaussian Mixture Models (GMM) classifiers. They described significant differences in classifier performances for detecting depression based on gender: 81-87% for males, and 72-79% for females.

With emphasis on vocal features for the identification of depression, Hönig et al. (2014) used automatic feature selection to study 34 features: spectral (MFCCs, formants F1 to F4), prosodic (pitch, energy, duration, rhythm) and vocal quality or phonetic features (jitter, shimmer, raw jitter, raw shimmer, logarithm harmonics-to-noise ratio, spectral harmonicity and spectral tilt). In agreement with findings from Low et al. (2011), they reported a slightly higher correlation in males (r = 0.39 males *vs*. 0.36 females). This suggests that clinical depression can possibly lead to more significant changes in vocal features in men than in women.

Similarly, Jiang et al. (2017) also noticed gender differences in classifier performances, with superior results in males. In a sample of 170 subjects, they investigated the discriminative power of three classifiers for the detection of depression: SVM, GMM and k-nearest neighbors (kNN). SVM achieved the best results in that study, with accuracy of 80.30% (sens.: 75%; spec.: 85.29%) for males, and 75.96% (sens.: 77.36%; spec.: 74.51%) for females.

Adversely, Higuchi et al. (2018) analyzed pitch (F0), spectral centroid and five attributes of MFCC using polytomous logistic regression for the classification of depression, bipolar disorder and healthy controls. They did not find any difference between genders. Additionally, they reported an overall accuracy of 90.79%, the highest among the studies revised for this work.

Another attribute of study design that might influence the performance of an automated instrument is the type of speech task. Spontaneous speech, e.g., social interactions or interviews tend to yield higher classification performances than reading tasks. This finding suggests that spontaneous speech provides more acoustic variability, improving the recognition of depression (Alghowinem, Goecke, Wagner, & Epps, 2013; Jiang et al., 2017). Moreover, it is likely that depressed individuals can suppress their emotional state during reading tasks, because of the unimportant nature of the read content or their concentration on reading, or even both (Mitra & Shriberg, 2015).

The rest of this work is divided as follows: Section 2 briefly discusses studies related to the identification of depression using voice. Section 2 describes in detail the implementation of a proposed voice-based instrument for the detection of depression. In Section 3 we present and discuss our results. Section 4 states our conclusion and suggestions for future work on this subject.

## 2. Methods

For this study 33 volunteers over 18 years old from both genders were selected and separated into one of the following groups:

▪ Control group: 11 healthy participants (6 males) were selected through the Self-Reporting Questionnaire (SRQ-20) screening for common mental disorders (Gonçalves, Stein, & Kapczinski, 2008; K. O. B. Santos, Araújo, Pinhoa, & Silva, 2010);
▪ Depression group: 22 patients with previous diagnosis of major depressive disorder (17 males), in conformity with Hamilton Depression Rating Scale – HAM-D 17 (Hamilton, 1960).

All individuals from the depression group fulfilled DSM-5 diagnostic criteria for major depressive disorder and were diagnosed by an independent professional prior to this study. Data for this group was collected in outpatient settings and psychiatric wards in Hospital das Clínicas, Federal University of Pernambuco, and in Hospital Ulysses Pernambucano, both in Recife, Northeast Brazil. Participants with coexistent neurological disorders or who made professional use of their voices were excluded. The use of validated psychometric scales aimed to verify previous diagnostic consistency and assess clinical severity. All participants have given written consent, and this study was conducted only after approval of a local Research Ethical Board. Table 1 provides a summary of mean age and scale scores for both groups.

**TABLE 1.**
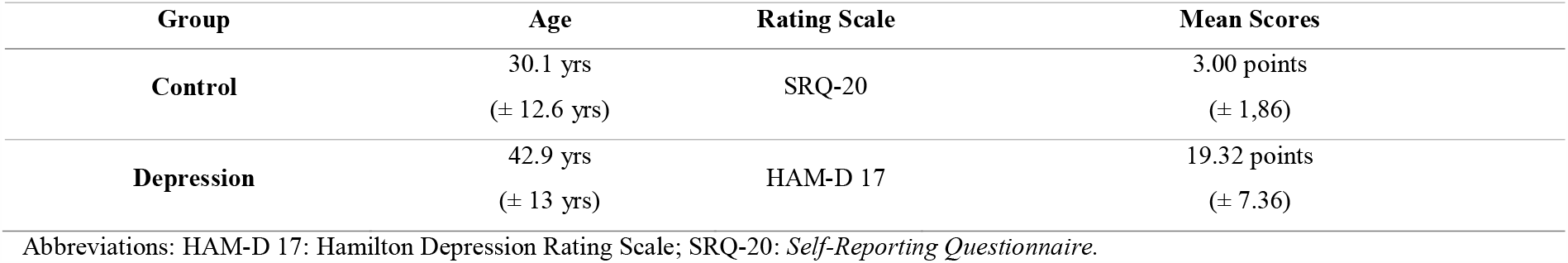
Mean age and rating scale scores

For the control group the SRQ-20 cutoff score was 6/7 (K. O. B. Santos et al., 2010), and for the depression group the eligibility criterion was HAM-D score above 7. Consequently, patients suffering from mild to severe depression were included.

### Acquisitions of voice samples

We used a Tascam™ 16-bit linear PCM recorder at 44.1KHz sampling rate, in WAV format, without compression. Audio acquisitions were made during an interview with a psychiatrist in naturalistic settings, i.e., patients from the depression group were recorded during a routine medical evaluation in an outpatient office or hospital ward. Recordings for the control group were made in different environments (e.g. offices, classrooms, gyms). After each interview, the clinician applied an appropriate rating scale (SRQ-20 or HAM-D 17) to verify diagnostic suitability. No duration limit was set for the recordings. As conversations were thoroughly recorded, voices from the clinician and possible third parties were also acquired and needed to be further removed. The total time recorded was 425.1 minutes (7.09 hours). Figure 1 summarizes the process of data acquisition.

**Fig. 1.**
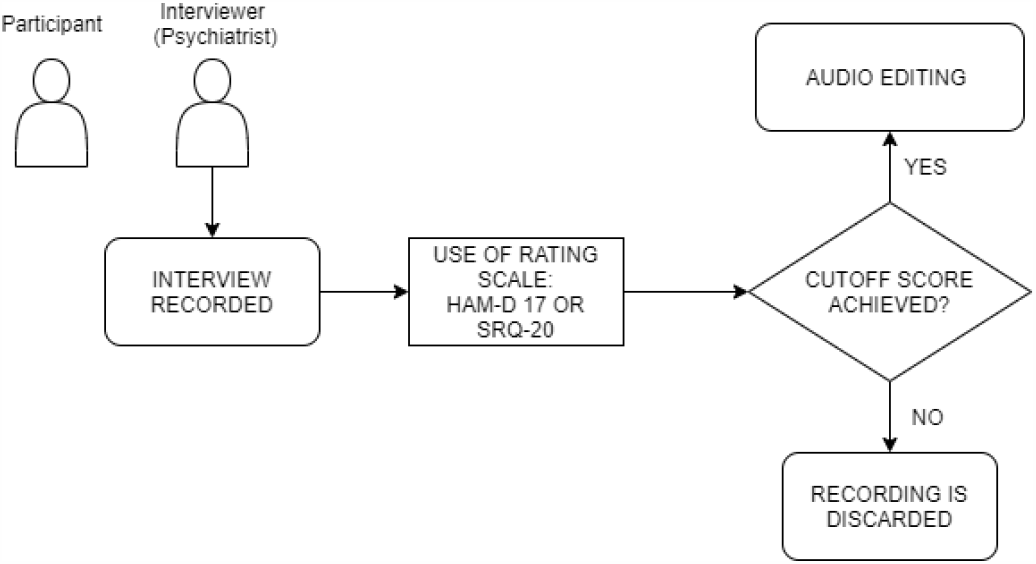
Block diagram of voice acquisition.

### Audio editing

After voice acquisition, we used Audacity™ audio software to remove voice signals from the interviewer and any potential companion. The edition process was manually made, and yielded 271 minutes of voice signals from participants (4.52h) as follows: 96.9 minutes for the control group, and 174.1 minutes for the depression group. Table 2 provides detailed information for both groups.

**TABLE 2.** Recording duration after audio editing

| Group | Number of participants | Total recording duration | Mean recording duration |
| --- | --- | --- | --- |
| Control | 11 | 5816s | 528s (8.8 min) |
|  |  | (96.9 min) | ± 138.7s |
| Depression | 22 | 10444s | 467,5s (7.8 min) |
|  |  | (174.1 min) | ± 213.3s |

### Feature extraction

All recordings were submitted to vocal feature extraction on GNU Octave™, a free open-source signal-processing software. We used rectangular windows, frame length of 10s with 50% overlap. As raw audio data was used, no filtering process was applied. During this stage, we extracted the following 33 features: skewness; kurtosis; zero crossing rates; slope sign changes; variance; standard deviation; mean absolute value; logarithm detector; root mean square; average amplitude change; difference absolute deviation; integrated absolute value; mean logarithm kernel; simple square integral; mean value; third, fourth and fifth moments; maximum amplitude; power spectrum ratio; peak frequency; mean power; mean frequency; median frequency; total power; variance of central frequency; first, second and third spectral moments; Hjorth parameter activity, mobility and complexity; and waveform length.

The choice of the above features relies on their accurate representation of input signals to computational models, because decision making process in machine learning does not depend on human interpretation. Furthermore, these features have already been successfully used for representing other signal types, such as electroencephalography.

### Classification

Both classes were balanced by adding artificial instances on Weka™ artificial intelligence environment. This step is important because it avoids computational biases towards the class with more representativeness, in this case the depression class. Features were submitted to experiments with the following ML algorithms on Weka™: multilayer perceptron (MLP), logistic regression, random forest (RF), decision trees, Bayes net, Naïve Bayes, and SVM with different kernels (linear, polynomial kernel, radial basis function or RBF, PUK, and normalized polynomial kernel). All experiments were performed using 10-fold cross-validation. Figure 2 summarizes the steps of our proposed solution.

**Fig. 2.**
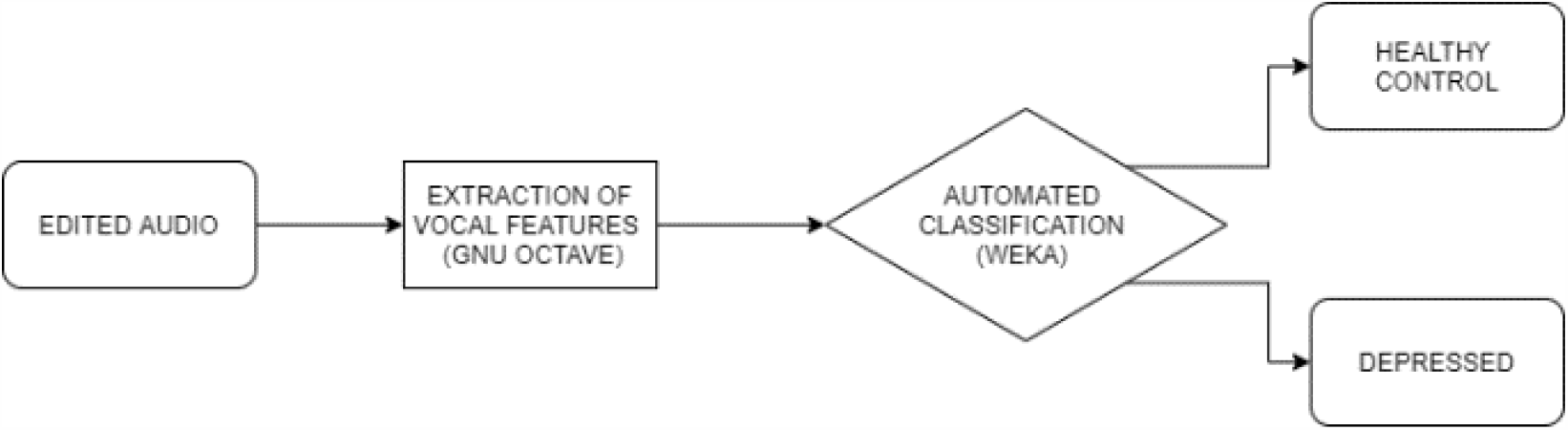
Block diagram of proposed solution.

## 3. Results

Experiments were initially made under default settings on Weka™. After this, we tested different setups for all algorithms with adjustable settings (MLP; polynomial kernel and normalized polynomial kernel SVM, SVM PUK kernel, and random forest). Table 3 describes in detail our best results for each ML model.

**TABLE 3.** Classification performance for machine learning algorithms – control *vs*. depression

| Model | Accuracy | Kappa | Sensibility | Specificity |
| --- | --- | --- | --- | --- |
| **MLP** | 81.84% | 0.6367 | 81.8% | 81.8% |
| **Random Forest** | 84.91% | 0.6982 | 84.9% | 85.0% |
| **Decision Trees** | 74.58% | 0.4916 | 74.6% | 74.6% |
| **Logistic Regression** | 68.69% | 0.3737 | 68.7% | 69.8% |
| **SVM linear** | 71.87% | 0.4373 | 71.9% | 73.5% |
| **SVM 2-Degree Polynomial** | 82.35% | 0.6471 | 82.4% | 82.9% |
| **SVM 3-Degree Polynomial** | 85.93% | 0.7186 | 85.9% | 86.1% |
| **SVM PUK** | **89.14%** | **0.7828** | **89.1%** | **89.2%** |
| **SVM normalized Polynomial** | 87.19% | 0.7438 | 87.2% | 87.2% |
| **SVM RBF** | 67.69% | 0.3539 | 67.7% | 72.0% |
| **Bayes Net** | 67.09% | 0.3418 | 67.1% | 67.7% |
| **Naive Bayes** | 60.01% | 0.2001 | 60.0% | 64.4% |

Abbreviations: MLP: *Multilayer Perceptron*; PUK: *Pearson Universal VII Kernel*; RBF: *Radial Basis Function*; SVM: *Support Vector Machines*.

## 4. Discussion

Through analysis of Table 3 above we notice that classification accuracy varied significantly for SVM (67.69%-89.14%), depending on the kernel used. SVM PUK kernel provided the highest accuracy (89.14%) among all classifiers in this study. The confusion matrix for this kernel is shown in Table 4. SVM normalized polynomial kernel and polynomial kernel achieved accuracy of 87.19% and 85.93% respectively. The greatest performances from different SVM kernels in this dataset supports findings from previous studies in the literature. This suggests the possible superiority of this algorithm for automated classification tasks using vocal acoustic features.

**TABLE 4.** Confusion matrix for the model with the highest performance (SVM PUK)

|  | Classified as<br>Control | Classified as depression |
| --- | --- | --- |
| Control | 88.25% | 11.75% |
| Depression | 9.97% | 90.03% |

As mentioned earlier, except for the work of Higuchi et al. (2018), we achieved better results than other previously published studies. Our work provided high classification accuracy both for depressed and healthy individuals. However, it is important to note that our small sample size may limit statistical interpretations. Factors that may influence vocal acoustic properties, such as smoking history and pharmacotherapy, were not controlled and represent a limitation. In a prospective study, we aim to control possible confounders and repeat the same experiments with more participants.

## 5. Conclusion

Current psychiatric diagnosis still lacks objective biomarkers, and relies mostly on specialist opinion based on diagnostic manuals. Nevertheless, such diagnostic systems have been heavily criticized due to their absence of correlation with the neurobiology and etiopathogenesis of mental disorders. Among these, depression presents with vocal acoustic alterations that may be used as objective parameters for the identification of this disorder.

Therefore, this study focused on the development of an auxiliary instrument for the diagnosis of depressive disorders. To this end, we extracted vocal acoustic features and performed experiments using different automated classification techniques. Some of the most widely used classifiers were also tested in this work. Our results demonstrate the viability of a machine learning tool for the detection and even screening of major depressive disorder in a cost-effective and non-invasive manner. In future studies we intend to perform the same experiments in a larger sample, as well as with gender-based datasets. We would like to assess if depression unequally affects vocal acoustic properties from men and women and how such differences influence the performance of automated classifiers.

## Data Availability

Data will be available when solicited

## Acknowledgements

The authors are grateful to the Brazilian research agency CNPq, for the partial financial support of this research.

## Compliance with Ethical Standards

This study was partially funded by the Brazilian research agency CNPq.

All procedures performed in studies involving human participants were in accordance with the ethical standards of the institutional and/or national research committee and with the 1964 Helsinki declaration and its later amendments or comparable ethical standards.

